# SARS-CoV-2 genomic surveillance from community-distributed rapid antigen tests

**DOI:** 10.1101/2024.08.12.24311680

**Authors:** Isla E. Emmen, William C. Vuyk, Andrew J. Lail, Sydney Wolf, Eli J. O’Connor, Rhea Dalvie, Maansi Bhasin, Aanya Virdi, Caroline White, Nura R. Hassan, Alex Richardson, Grace VanSleet, Andrea Weiler, Savannah Rounds-Dunn, Kenneth Van Horn, Marc Gartler, Jane Jorgenson, Michael Spelman, Sean Ottosen, Nicholas R. Minor, Nancy Wilson, Thomas C. Friedrich, David H. O’Connor

**Affiliations:** University of Wisconsin-Madison School of Medicine and Public Health, Madison, Wisconsin, USA; Madison West High School; University of Wisconsin-Madison, Wisconsin National Primate Research Center, Madison; Public Health Madison Dane County, Madison; Madison Public Library, Madison; University of Wisconsin-Madison School of Veterinary Medicine, Madison

**Keywords:** SARS-CoV-2, Public Health, Wisconsin, RNA, COVID-19

## Abstract

In the United States, SARS-CoV-2 genomic surveillance initially relied almost entirely on residual diagnostic specimens from nucleic acid amplification-based tests (NAATs). The use of NAATs waned after the end of the COVID-19 Public Health Emergency. We partnered with local- and state-level public health agencies and the Dane County Public Library System to continue genomic surveillance by obtaining SARS-CoV-2 genome sequences from freely available community rapid antigen tests (RATs). From August 15, 2023 to February 29, 2024 we received 227 tests, from which we generated 127 sequences with >10x depth of coverage for ≥90% of the genome. In a subset of tests, lower Ct values correlated with sequence success. Our results demonstrate that collecting and sequencing from RATs in partnership with community sites is a practical approach for sustaining SARS-CoV-2 genomic surveillance.

## Introduction

Genomic surveillance is a powerful tool that can inform public health responses to disease outbreaks (1). During the COVID-19 pandemic, genomic surveillance data were used to identify variants of concern, investigate patterns of transmission, and develop effective vaccines (2–4).

Genomic surveillance requires large, representative sets of samples. Initially, nucleic acid amplification tests (NAATs) were the gold standard for detecting SARS-CoV-2 infection (5). Residual nasal swab samples leftover from NAAT testing were provided to laboratories for viral sequencing through contracts with companies and clinics performing NAATs. After the COVID-19 public health emergency ended on May 11, 2023, NAAT testing in clinical and public health facilities declined precipitously as government subsidies for the performance of NAATs ended (6). During 2020, an average of ∼587,975 NAATs were performed weekly in the US. By 2023, this decreased to ∼96,215 tests (7). Subsequently, the primary source of samples for genomic surveillance was greatly diminished.

The CDC now recommends at-home COVID-19 rapid antigen tests (RATs) for those without access to NAATs (8). RATs are cheaper than NAATs, provide faster results, and do not require trained personnel (9). RATs usually require swabbing the insides of both nostrils, placing the swab into an inactivation buffer, and applying the buffer onto a lateral flow test strip. If SARS-CoV-2 antigen is present, a colorimetric test line indicates positivity (10). As of this writing in July 2024, there are 38 over-the-counter antigen test products authorized by the FDA in the US (11).

Multiple groups have investigated RATs as source material for SARS-CoV-2 genomic surveillance. SARS-CoV-2 viral nucleic acids can be recovered from antigen tests and sequenced (12–16), allowing for circulating lineages to be tracked.

We hypothesized that people would be willing to send in SARS-CoV-2-positive RATs for genomic surveillance if the process was sufficiently easy. To test this hypothesis, we created a system for individuals to anonymously submit positive RATs. We partnered with public libraries and a public health agency in Dane County, Wisconsin USA to bundle mail-in research packets along with freely available RATs. Upon testing positive for SARS-CoV-2, community members could mail positive tests to our laboratory for sequencing.

## Materials and Methods

### Ethics statement

The University of Wisconsin IRB determined this project was human research exempt because participants were anonymous and self-identified. A secure website and database were created using Node JS to collect the barcode, the date, and the location when a user scanned a randomly generated unique QR code. We assumed these data were a reasonable proxy for RAT assay date and location. The location of the scan was automatically converted to a census block group on the users’ machines before submission to our database, so the actual location of each submission was not known to the study team. A census block group contains between 250 and 550 housing units (17).

### Collection of Rapid Antigen Tests

In Wisconsin, the Dane County Public Library System and Public Health Madison Dane County (PHMDC) distributed RATs from the US national stockpile to the public free of charge. We partnered with nine libraries and two distribution sites through PHMDC. Four of the libraries were in Madison and five were in more rural areas of Dane County (Figure 1).

**Figure 1.**
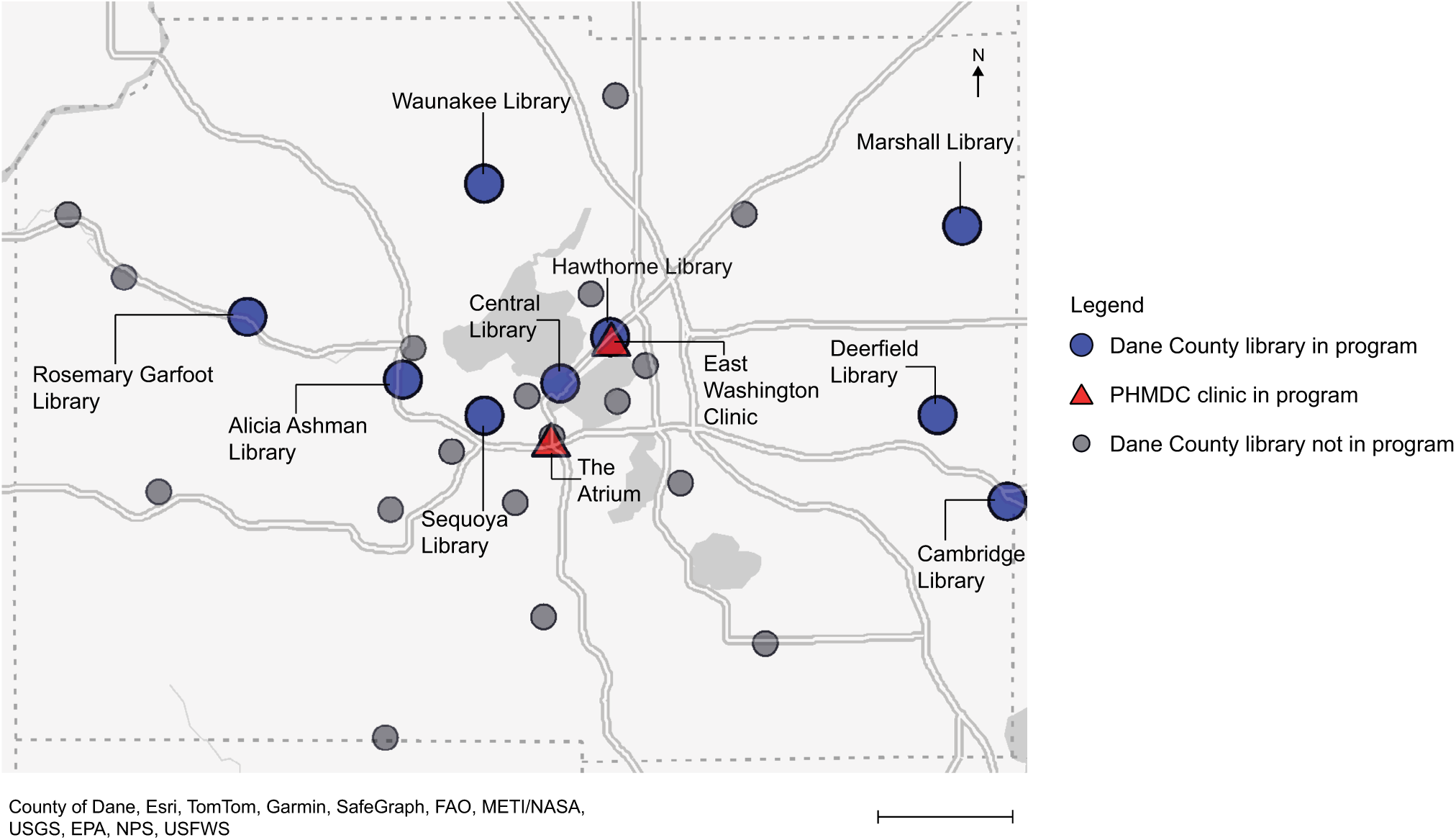
Map of Dane County, WI showing the locations of all Dane County libraries and Public Health Madison Dane County (PHMDC) clinics. The scale bar represents 10 miles. Nine of the Dane County libraries and two PHMDC sites were involved in handing out research packets along with SARS-CoV-2 rapid antigen tests to patrons, which allowed willing participants to send their positive tests to our laboratory to be sequenced. The nine libraries that were active in this program are shown as blue circles. The two PHMDC clinics that also distributed research packets and tests are shown as red triangles. The gray circles depict the locations of other libraries in the Dane County Library System that were not involved in the program.

We designed a packet of materials to attach to each RAT to enable positive test collection (Figure 2). This packet included a bubble mailer with a business reply mail shipping label, a zip-lock bag with a unique QR barcode inside which was placed inside the mailer, and an instructional flyer affixed to the outside. The flyer had instructions in both English and Spanish, describing the study and providing instructions to participate (Appendix Figure 1).

**Figure 2.**
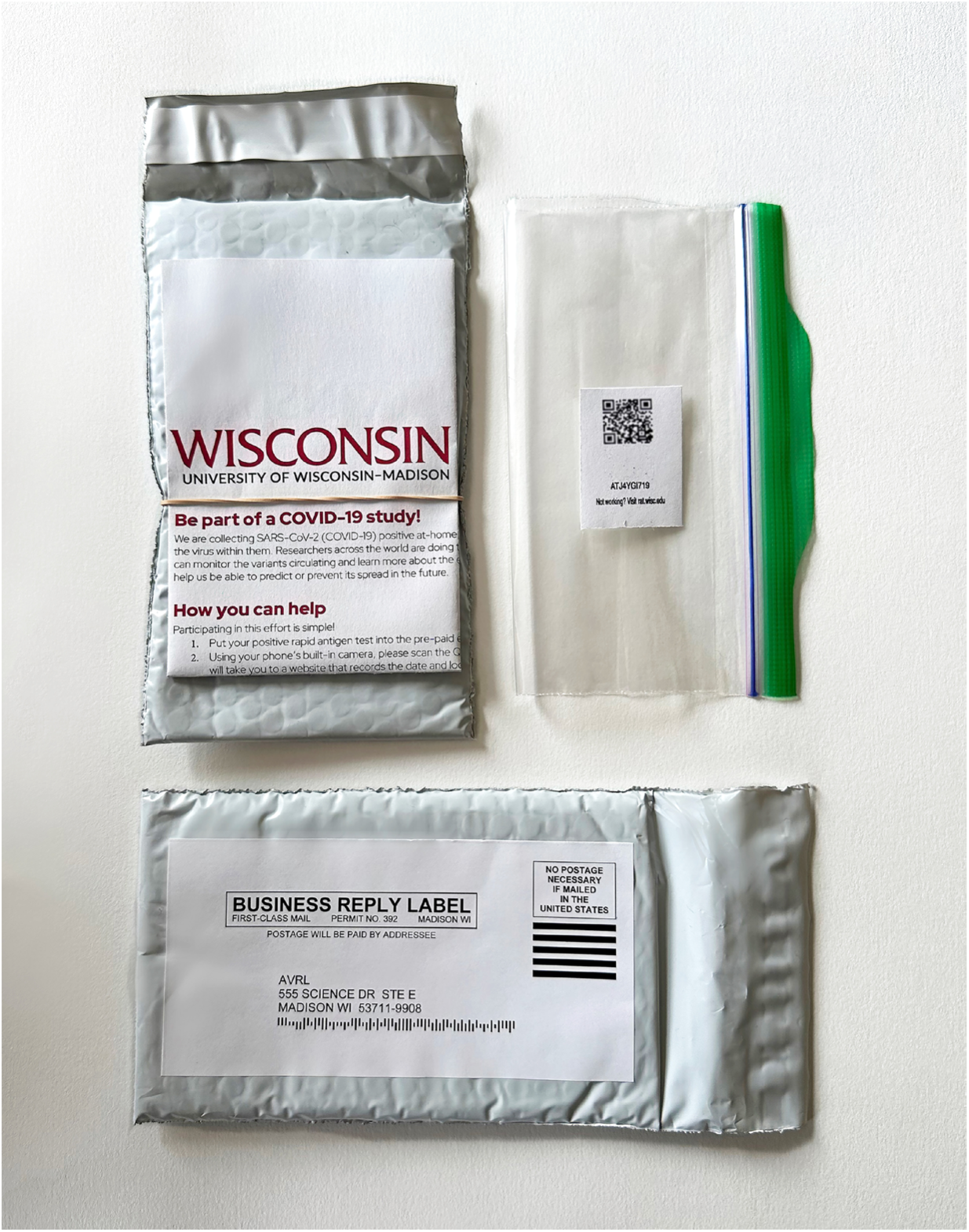
An image of the packet that was attached to SARS-CoV-2 rapid antigen test boxes, enabling participants to send their positive tests to the lab through the USPS. In the upper left is a folded flyer attached to an envelope. The flyer explains the goal of our study and how to participate in both English and Spanish. The upper right shows a zip-lock bag with a QR code placed inside, which participants scanned to document the date and location of their test. The location of the scanned QR code was immediately converted to the census block group of the scan and stored in our secure database. The bottom shows a business-reply shipping label pasted onto the envelope, allowing participants to drop their sealed envelope at a post office drop box to be sent to our lab.

If a RAT was positive, participants could volunteer to submit the test to our program. They were directed to scan the unique QR code on an internet-connected device and drop their sealed envelope with the positive test in any post office mailbox. The inactivation buffer in a RAT inactivates SARS-CoV-2, rendering the tests non-biohazardous and therefore safe to send through the mail (18).

Upon arrival at the lab, the QR code was scanned to record the date of receipt. Each RAT was stored at -80°C until processing. The majority of RATs we received were BinaxNOW^TM^ COVID-19 Antigen Self Tests (Abbott, https://www.abbott.com) or iHealth COVID-19 Antigen Rapid Tests (iHealth Labs Inc., https://ihealthlabs.com).

### Extraction of Nucleic Acids

We developed our approach to extract nucleic acids from used RATs based in part on methods originally described by Martin et al. (12).

Tests were thawed and opened to retrieve the testing strip, which was placed into a clean 5 mL Sarstedt freezer tube (Sarstedt, https://www.sarstedt.com/en). Some tests also included a nasal swab; these were also placed in the freezer tube.

We added 800 μL of Viral Transport Medium (VTM, Rocky Mountain Biologicals, LLC, <u>https://</u> <u>rmbio.com</u>), and the tube was incubated at room temperature for ten minutes on a Hulamixer (ThermoFisher Scientific, https://www.thermofisher.com). 500 μL were transferred to a clean 1.5 mL tube, and 5 μL of Dynabeads Wastewater Virus Enrichment Beads (ThermoFisher Scientific) were added. Subsequently, we followed the manufacturer’s protocol for the MagMAX™ Wastewater Ultra Nucleic Acid Isolation Kit with Virus Enrichment (ThermoFisher Scientific). Samples were isolated on a Kingfisher Apex instrument (ThermoFisher Scientific) following the manufacturer’s protocol (MagMAX_Wastewater_DUO96.bdz).

Following isolation, the samples were treated with Turbo^TM^ DNase (ThermoFisher Scientific) according to the manufacturer’s protocol. After DNAse treatment, samples were cleaned using the RNA Clean and Concentrator-5 kit (Zymo Research, https://www.zymoresearch.com) following the manufacturer’s protocol, skipping the in-column DNase II Treatment.

### RT-qPCR

We selected a random subset of 75 samples to investigate trends between the RT-qPCR Ct and sequencing quality. We quantified SARS-CoV-2 vRNA using the CDC N1 Taqman assay (19). Details for how these RT-qPCR assays were set up are available in the Appendix.

### RT-PCR and Sequencing

PCR amplicons were generated using the QIAseq DIRECT SARS-CoV-2 Kit with Booster and Enhancer (QIAGEN) according to the manufacturer’s instructions. Indexed samples were normalized to 4 nM and pooled together. The pool was diluted to a concentration of 8 pM and run using 2x150 Miseq Reagent Kits v2 (Illumina, https://www.illumina.com) on a MiSeq instrument (Illumina).

### Sequencing Analysis

Raw sequencing reads were quality-checked, aligned to the Wuhan-1 SARS-CoV-2 reference (MN908947.3), and variant-called using the open-source *viralrecon* pipeline from the *nf-core* project (20–22). The minimum frequency threshold for variant-calling was set to 0.01. Further details for how we ran *viralrecon*, alongside the custom R scripts we used to generate figures, are available in our <u>Library-Rapid-Antigen-Test-Manuscript</u> Github repository.

### Statistical Analysis

The effect of Ct and length of transit time was compared between samples that passed our sequencing quality threshold of ≥90% coverage at >10x depth and those that failed, using an unpaired two-tailed t-test (Prism v10.1.0).

We compared the identities of SARS-CoV-2 lineages detected in our RAT-associated sequences with surveillance data from the Wisconsin State Lab of Hygiene’s (WSLH) SARS-CoV-2 Genomic Dashboard (23). We analyzed data from August 28th, 2023 to February 25th, 2024, dividing our passing sequences into two-week intervals based on test scan dates. Only participant-scanned tests were included in this analysis. We assigned Pango lineages to our sequences using Nextclade v3.5.0 (24). For each two-week period we identified the two most prevalent lineage groups, which are based on Nextstrain clades, in the WSLH wastewater surveillance data and determined how often our RAT program also detected these two most prevalent lineages.

## Results

### Test Collection

Between August 15th, 2023, and February 29th, 2024, we supplied nine Dane County libraries and two public health clinics with 7,775 research packets to attach to SARS-CoV-2 RATs distributed to patrons. 223 of these packets (2.9%) were mailed to our lab. Some packets contained multiple tests, resulting in 227 tests for analysis. The return rates varied by month (Table 1), but the mean number received each month was 32 ± 10.

**Table 1.**
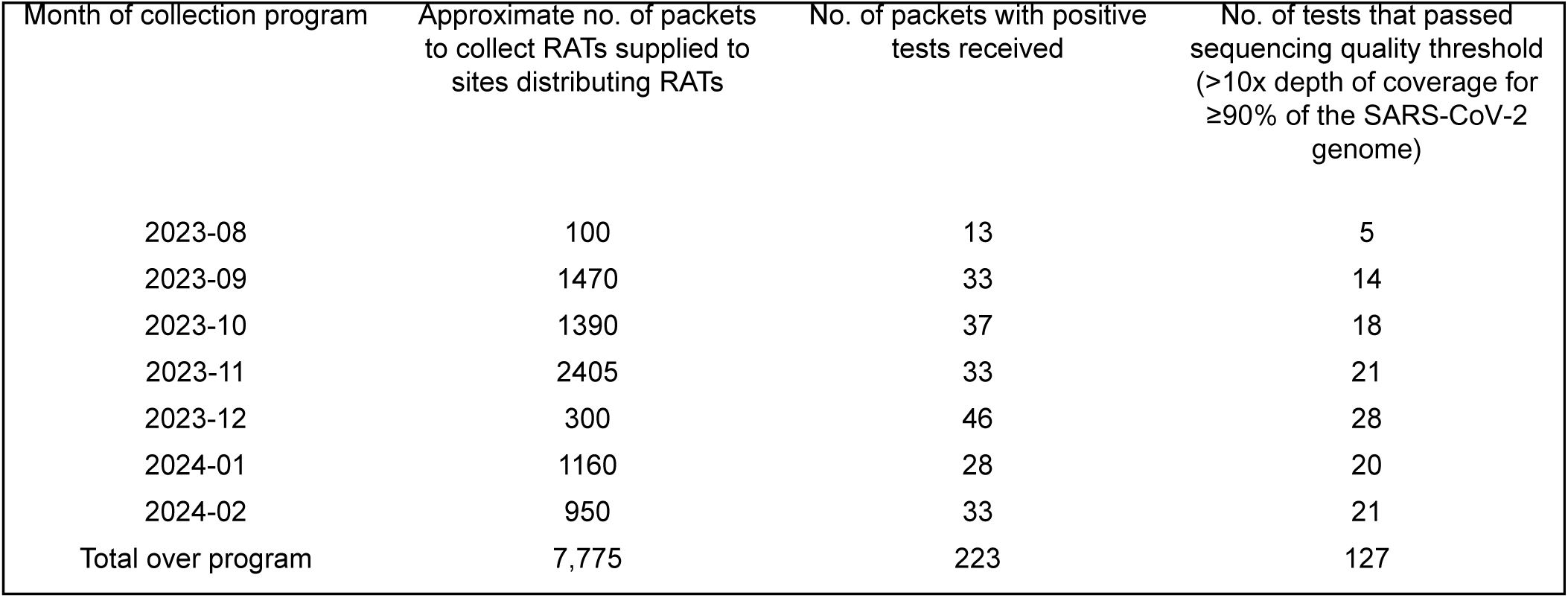
Monthly rate of materials supplied to collect positive rapid antigen tests, positive tests received in our lab, and tests that passed our sequencing quality threshold.

Some tests arrived without the barcode or with a barcode that had never been scanned, resulting in no associated metadata. Of the 223 research packets received, 170 were properly associated with time and location metadata. Of these 170 samples, one was scanned in Sauk County, Wisconsin (adjacent to Dane County), and the rest in Dane County, Wisconsin.

### Sequencing Quality

We sequenced SARS-CoV-2 from all 227 RATs. We considered a sequence with genome coverage ≥ 90% at a depth of coverage > 10x to be a “passing” sequence. 128 of the 227 RAT-derived sequences passed, a success rate of 56%. Sequencing metrics for each sample are provided in Appendix Table 1.

Next, we evaluated whether SARS-CoV-2 vRNA concentration or transit time correlated with successful sequencing. 75 samples were randomly selected for semi-quantitative RT-qPCR. 15 of these had no detectable amplification of the N1 target. We obtained passing sequences for samples with Ct values up to 35.4. The mean Ct for samples that passed was 31.7 and the mean Ct for those that failed was 35.3, a significant difference (Figure 3A; unpaired t-test, two-tailed p < 0.0001, df = 59).

**Figure 3.**
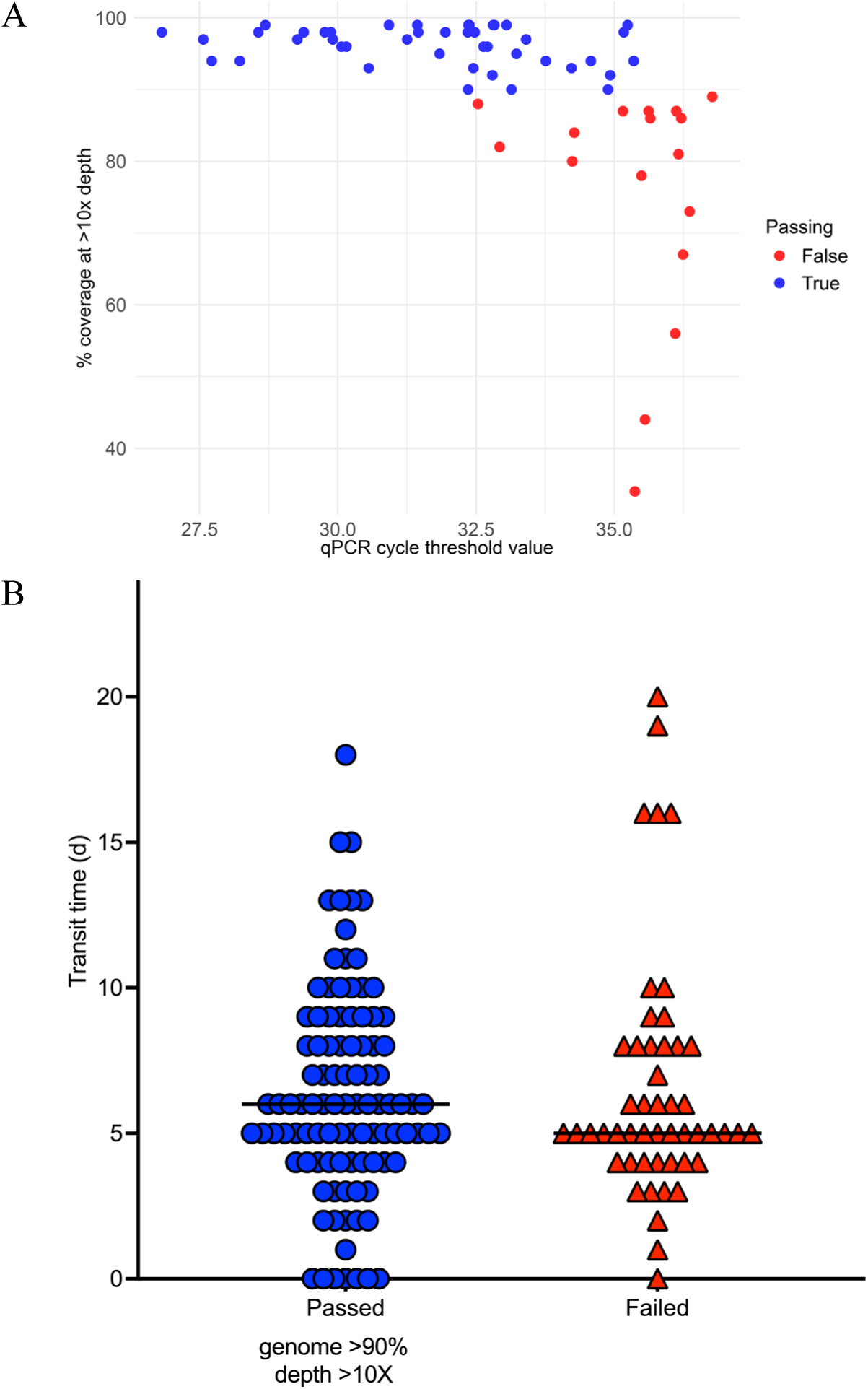
The amount of viral material present correlated with our ability to sequence rapid antigen test (RAT) samples, but time en route did not. A) A scatterplot comparing the Cycle Threshold (Ct) value obtained through reverse transcriptase quantitative PCR (RT-qPCR) to the percent coverage of the SARS-CoV-2 genome at >10x depth of coverage obtained from each RAT sample. For a sequence to pass our sequencing quality threshold, it must cover ≥90% of the SARS-CoV-2 genome at >10x depth. Samples that passed this threshold are colored blue, and those that did not are colored red. The mean Ct for samples that passed was 31.78 and the mean Ct for those that failed was 35.34 (unpaired t-test, two-tailed p < 0.0001, df = 59). Samples with lower Ct values were correlated with higher SARS-CoV-2 coverage. B) This scatterplot shows the time (in days) between when the QR code was scanned by a participant and when our lab received the sample, referred to as the transit time. These tests are separated into those that passed the sequencing quality threshold (blue circles) as defined above and those that failed (red triangles). The horizontal black line is the median value for each group. The mean transit time for the samples that passed was 6.6 days and the mean transit time for samples that failed was 6.3 days. There was no significant difference between these groups (unpaired t-test, p = 0.58).

The time between a test being scanned by the participant and our receiving it (i.e., the transit time) ranged from one to twenty days. Figure 3B illustrates the relationship between transit time and sequencing success. The mean transit time for the samples that passed was 6.6 days, compared to 6.3 days for samples that failed (unpaired two tailed t-test, p = 0.58).

### Tracking SARS-CoV-2 Lineages

We used Nextclade v3.5.0 (24) to determine the Pango lineage of each successfully sequenced sample. Figure 4 shows SARS-CoV-2 lineages detected by week based on participant scan date. From August to November 2023, the majority of lineages were assigned to the XBB* clade. Beginning in December 2023, we observed a shift to the lineage JN.1*, which predominated in February 2024.

**Figure 4.**
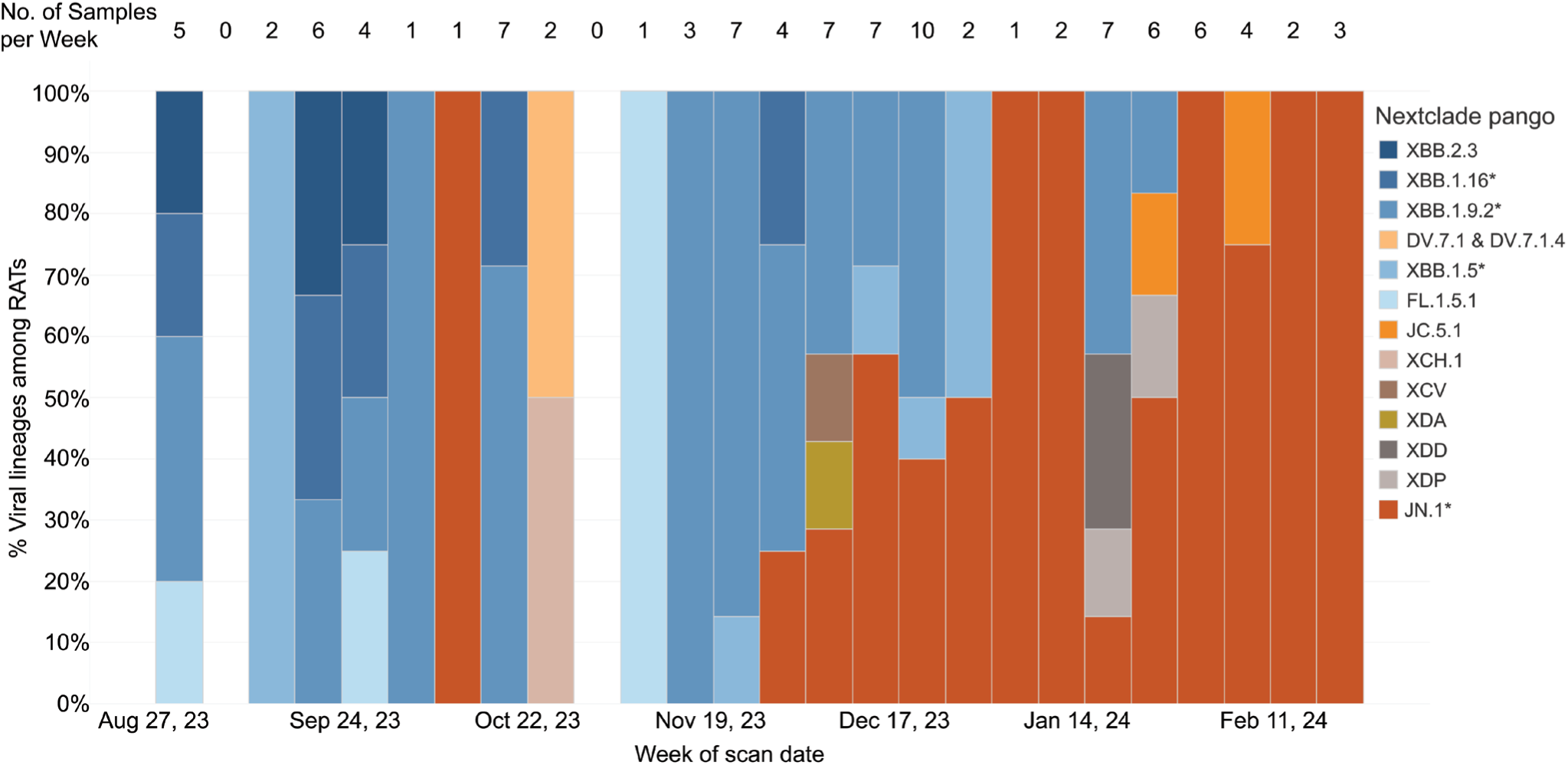
Rapid antigen test lineages determined for each week. This chart shows the proportion of SARS-CoV-2 lineages by week that passed QC (≥90% of the SARS-CoV-2 genome at >10x depth). The date used was the date the QR code was scanned by the participant. RATs that were not scanned were excluded from this analysis. The number of samples included in each week’s proportion is shown as a number above the bar. From August to mid-November, the most common lineages in our samples fell under XBB.1.5*, XBB.1.9.2*, XBB.1.16*, and XBB.2.3. Beginning in early December, we began to see an increase in the number of samples belonging to the lineage JN.1*. This lineage dominated RAT samples scanned in February 2024.

The identities of viral lineages in our RAT-derived sequences were concordant with statewide trends in lineages detected via wastewater surveillance, as summarized on the WSLH SARS-CoV-2 Wastewater Genomic Dashboard (23) (Table 2). Our program detected the dominant wastewater lineage in 12 of 13 two-week reporting periods and the second-most prevalent lineage in 7 of 13 periods. Concordance with wastewater surveillance data indicates that RAT-based surveillance can detect common circulating lineages. Moreover, RAT-based surveillance resulted in six of the earliest documented cases of a lineage in Wisconsin in Genbank and GISAID: JN.1.1, JN.1.2, XDD, XDA, XDP, and XDE (Table 3).

**Table 2.**
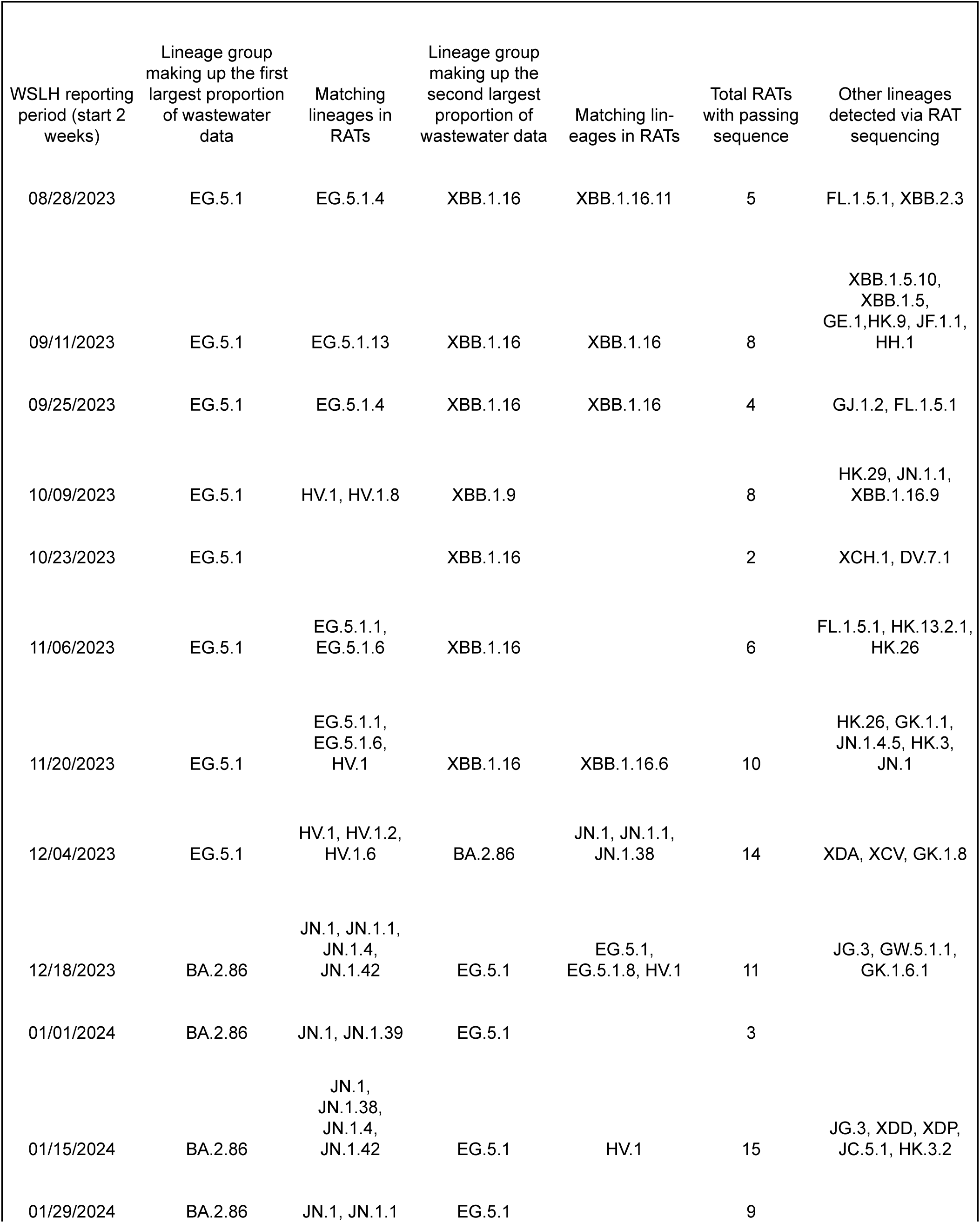
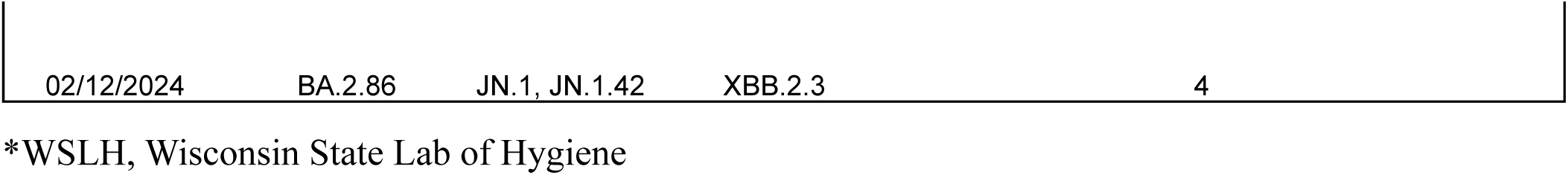
List of lineages of our successful rapid antigen test sequences that corresponded to the two most prevalent lineage groups in the wastewater signal in the state of Wisconsin for each two week period.

**Table 3.** Rapid antigen test samples that are the earliest recorded example of their respective Pango lineage in the state of Wisconsin according to data submitted to GISAID and Genbank as of April 18, 2024.

| Genbank accession number | Scanned date of test | Pango lineage |
| --- | --- | --- |
| PP761647 | 10/14/23 | JN.1.1 |
| PP747716 | 12/4/23 | XDA |
| PP747739 | 12/21/23 | JN.1.2 |
| PP747779 | 12/22/23 | XDE |
| PP747696 | 1/17/24 | XDD |
| PP747750 | 1/24/24 | XDP |

## Discussion

Genomic surveillance has been crucial in tracking SARS-CoV-2 evolution during the COVID-19 pandemic (25). With most individuals now using RATs instead of NAATs, we sought to evaluate a community genomic surveillance program predicated on the voluntary mailing of positive RATs.

Despite the common narrative that the public is disinterested in COVID-19, we observed surprisingly strong participation. Between August 12, 2023 and February 24, 2024, Dane County’s COVID-19 test positivity averaged 12.3% (7.8-16%) (26). If we assume an extreme case in which all the RATs distributed by our partners were used, we estimate that a quarter of all positive tests distributed with packets were returned to our laboratory for analysis. The true return rate is likely higher given that it is unlikely that all tests with packets were used.

The transit time during which RATs sat in uncontrolled (ambient) conditions had a negligible impact on overall sequencing success (Figure 3b). Other studies have demonstrated that extraction of nucleic acids is possible from RATs stored at room temperature for long periods of time (13,15); one study generated 75.2% genome coverage from a RAT stored at room temperature for 3 months. We obtained a sequence with >10x coverage for ≥90% of the SARS-CoV-2 genome from a RAT that sat at uncontrolled temperatures for at least 17 days. Taken together, these results highlight that RATs stored at ambient temperature can be mailed from the point-of-testing to centralized labs for sequencing. 98% of the United States population is served by the United States Postal Service (27). The ability to self-collect samples for mail-in analysis could enable genome surveillance even in settings that are typically underserved by academic and clinical research.

SARS-CoV-2 lineages identified by our RAT surveillance program were similarly prevalent in Wisconsin’s statewide wastewater sequencing data (23). Notably, we also detected emerging lineages like JN.1 and rare variants like XDE (documented only 22 times in North America (28)). These findings demonstrate that RAT-based sequencing can effectively complement existing wastewater and NAAT surveillance methods.

An important limitation of our study is the reliance on self-reported data, which is less precise than clinical specimen metadata. Our metadata depends on the participant’s QR code scan to approximate the date and location of the test, which may reduce data accuracy. Another limitation is that ∼25% of envelopes arrived unscanned or without a barcode, resulting in no metadata. Our only communication with these participants is through the flyer provided with each packet, and some participants may only skim over the instructions, resulting in misinterpretations of the protocol. To reduce the frequency of unscanned tests, more comprehensive instructions could be provided along with simplified visuals to grab attention and more clearly communicate these directions.

Our program is reliant on freely available RATs. Currently, these tests are provided from the national government stockpile. The long-term sustainability of the programs that distribute these tests is unknown, which means this system for the collection and sequencing of RATs may not be sustainable long-term. While a similar program could be established with RATs purchased by community members (e.g., by partnering with pharmacies to put them at point-of-sale), this could significantly bias the results towards those who have the resources and motivation to purchase costly tests. Providing free tests to members of the community gives them a valuable tool to minimize their risk of COVID-19 transmission while also potentially resulting in more inclusive, representative genomic surveillance.

The program described here could act as a framework for the creation of more expansive genomic surveillance programs. Regulators in some countries have approved at-home RATs for other respiratory viruses including IAV and RSV (29–31), and these tests could be collected to set up surveillance programs for other viruses. As studies have demonstrated the possibility of recovering various respiratory viruses from COVID-19 RATs (14,32), the prevalence of respiratory viruses present in communities could also be estimated by collecting both positive and negative RATs from individuals who are symptomatic, creating an innovative additional method for assessing the spread of respiratory viruses in communities.

## Data Availability

The sequencing data generated in this study are available in the Sequence Read Archive (SRA) under the BioProject PRJNA1096364. The accession numbers to the sequences used in these analyses are available in Appendix Table 2.

## Data Availability

The sequencing data generated in this study are available in the Sequence Read Archive (SRA) under the BioProject PRJNA1096364. The accession numbers to the sequences used in these analyses are available in Appendix Table 2 of this manuscript.

https://github.com/dholab/Library-Rapid-Antigen-Test-Manuscript

## Acknowledgments

This work was supported by the Wisconsin Department of Health Services [435100-A24-ELCProjE] and the CDC [75D30122C15355]. Analysis of these data was made possible by the Center for High Throughput Computing’s High Performance Cluster at the University of Wisconsin-Madison. The chatbot Claude 3.5 sonnet was used to improve wording choices and to make the text more concise. The Wisconsin Department of Health Services provided RATs to libraries and the public, during and after the COVID-19 public emergency. We would like to thank the many patrons of Dane County Public Libraries and PHMDC who were kind enough to take time while they were sick to scan and send us their tests. Without these community members, this study would not be possible. We would also like to thank all the staff at the Dane County Public Libraries who helped us to implement this program. We would like to specifically thank Leah Fritsche, Erick Plumb, Matt Rahner, Samantha Seeman, and Elizabeth Clauss, who agreed to distribute these packets at their libraries. We would also like to thank Sam Petykowski, a high school student who volunteered his time to produce research packets for Alicia Ashman Library.

## Biographical Sketch

Isla Emmen is a Research Specialist performing infectious disease research at the David O’Connor Laboratory at the University of Wisconsin-Madison in the School of Medicine and Public Health. She received her Bachelor of Science at the University of Wisconsin-Madison in 2023.

