## Appendix for "SARS-CoV-2 genomic surveillance from community-distributed rapid antigen tests"

### RT-qPCR Methods:

To quantify SARS-CoV-2 RNA, we used the CDC N1 Taqman assay (1). The forward primer sequence was “GACCCCAAAATCAGCGAAAT,” the reverse primer sequence was “TCTGGTTACTGCCAGTTGAATCTG,” and the probe sequence was “ACCCCGCATTACGTTTGGTGGACC.” A 20 uL reaction was set up with 5 uL of Taqman Fast Virus 1-step Master Mix (ThermoFisher Scientific, https://www.thermofisher.com), 1.5 uL of SARS-COV2 RUO Primer/probe kit N1 (Integrated DNA Technologies, https://sg.idtdna.com), 8.5 uL of nuclease-free water, and 5 uL of sample RNA. The assay was run on a LightCycler 96 instrument (Roche, https://www.roche.com) with cycling conditions of 37°C for 2 min, 50°C for 15 minutes, 95°C for 2 minutes, and 50 cycles of 95°C for 3 seconds and 55°C for 30 seconds.

To confirm successful isolation of viral nucleic acid from the RAT substrate, each sample was also tested for the presence of human ribonuclease P nucleic acid. Only samples which tested positive for ribonuclease P were included in the analysis.

WISCONSIN
UNIVERSITY OF WISCONSIN–MADISON

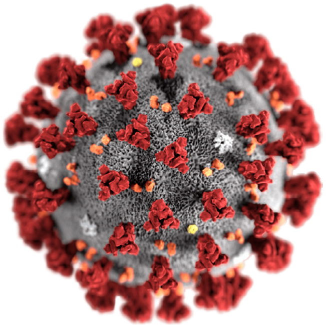

### Be part of a COVID-19 study!

We are collecting SARS-CoV-2 (COVID-19) positive at-home rapid antigen tests to identify the virus within them. Researchers across the world are doing this identification process so we can monitor the variants circulating and learn more about the evolution of COVID-19. This will help us be able to predict or prevent its spread in the future.

### How you can help

Participating in this effort is simple!

1. Put your positive rapid antigen test into the pre-paid envelope.
2. Using your phone's built-in camera, please scan the QR code in the Ziploc, which will take you to a website that records the date and location when you scan the code. Your phone's camera should already have the ability to recognize QR codes. You should not need to download an app for this.
3. If you have trouble scanning the QR code, you can visit http://rat.wisc.edu and type your code in manually.
4. Please place the barcode back inside the Ziploc before sealing the envelope.
5. Drop the sealed envelope into a mailbox. Thank you!

### We will protect your privacy

While we will not collect any personal information about you, scanning the QR code will record your location and the time you scanned the code, which is considered to be private information. These data will help us to determine when and where the virus was present in the community. These data will be stored in a secure database with access only for the scientists on the study. After we sequence your sample and determine the variant, we will upload the sequence information, date of scan, and in which county the QR code was scanned to national databases. These national databases (NCBI Virus and NCBI SRA) are a resource for all scientists doing COVID research and contain millions of sequences.

### More about this project

Throughout the COVID pandemic, the David O'Connor group at the University of Wisconsin has been sequencing COVID from nasal swabs to determine variants active in our community. As rapid antigen tests have become the main type of COVID test, we have developed methods to sequence the virus directly from these tests. Our objective is to continue sequencing samples in order to understand the variants present in the community. If you have questions about this project, please contact us at 608-890-0847. You can look up some of our work at: https://dho.pathology.wisc.edu/.

**Appendix Figure 1. Contents of the flyer attached to each rapid antigen test collection packet.** This flyer explains the goal of the study and provides instructions for those who want to participate. The flyer was printed double-sided and included an English translation and a Spanish translation (not shown). Please reach out to the corresponding author if you would like to see the Spanish version of this flyer.

**Appendix Table 1. Sequencing metrics for each rapid antigen test sample**

| Sample Name | % Coverage >10x | Input Reads | Mapped Reads |
|---|---|---|---|
| Lib RAT 1 CC | 89 | 292498 | 149451 |
| Lib RAT 2 CC | 99 | 429670 | 379054 |
| Lib RAT 3 CC | 87 | 906378 | 386605 |
| Lib RAT 4 CC | 99 | 751628 | 560350 |
| Lib RAT 5 CC | 96 | 638864 | 509354 |
| Lib RAT 6 CC | 73 | 939016 | 248200 |
| Lib RAT 7 CC | NA | NA | NA |
| Lib RAT 8 CC | 38 | 890258 | 246596 |
| Lib RAT 9 CC | NA | 2 | NA |
| Lib RAT 10 CC | 97 | 576444 | 363896 |
| Lib RAT 11 CC | 26 | 1031716 | 216466 |
| Lib RAT 12 CC | 69 | 739654 | 296681 |
| ATQC4UXFHL | 94 | 567612 | 469708 |
| ATGRM0X3NG | 99 | 518862 | 475772 |
| AT45CHNPMD | 89 | 814054 | 306016 |
| ATFFPBDGC7 | 99 | 901902 | 587391 |
| ATRUPA5DJ5 | 97 | 644592 | 553230 |
| ATKCPI0YMU | 98 | 121384 | 115142 |
| ATU2UEZGUU | 91 | 920378 | 687997 |
| AT25TA545F | 95 | 569574 | 516194 |
| ATAC3S9AX4 | 99 | 463772 | 426031 |
| AT1XJPLMT1 | 50 | 1835618 | 389978 |
| ATX2RLNE3M | 99 | 508792 | 451110 |
| AT9M50GXL5 | 98 | 565532 | 380139 |
| ATFY3C92IS | NA | 506 | 82 |
| ATBUK9BEKQ | 46 | 703636 | 118219 |
| AT1REAXVVZ | 91 | 720132 | 354056 |
| ATHXJM6OTS | 99 | 479808 | 413933 |

| | | | |
|---|---|---|---|
| No barcode 1 | NA | 788 | 734 |
| AT7V74R0RW | 99 | 412494 | 341780 |
| ATXPYXN5EQ | 88 | 842312 | 258262 |
| ATC50LYO64 | 5 | 1743894 | 369998 |
| ATILJWETL2 | NA | 700 | 622 |
| ATGCWPGJ2V | 9 | 1224 | 1116 |
| AT7ZH839N8 | 94 | 1170808 | 226765 |
| ATYFNYVCLA | 26 | 1772 | 1622 |
| AT2I45F27V | NA | 960 | 862 |
| ATHDDZ3Z8S | 83 | 830390 | 295767 |
| AT13LPB6FO | 46 | 1318052 | 214090 |
| ATL5VRS761 | 48 | 883866 | 154114 |
| ATZB3XY1WP | 79 | 722400 | 172355 |
| AT137YUSYR | NA | 1022 | 914 |
| ATF5O133LQ | 96 | 585228 | 198721 |
| ATRBJD97CM | 100 | 2942536 | 2511305 |
| AT6DRTU76H | 37 | 843298 | 133620 |
| ATUQ7230RN | 99 | 991114 | 276971 |
| AT5UTIINHL | 98 | 764184 | 171129 |
| AT0MKPPLPR | 68 | 668400 | 132895 |
| ATCLJ5EIQG | 99 | 448282 | 317652 |
| ATPCXFPKEX | 59 | 765374 | 120615 |
| AT8X5RUNJL | 97 | 484554 | 290255 |
| AT5R2HNUD3 | 57 | 1295960 | 196832 |
| AT2HXD2MB5 | 1 | 1142774 | 151226 |
| AT4CHNRA3T | 3 | 930698 | 101019 |
| ATCOXC4GN2 | 8 | 1405954 | 226976 |
| AT06HSAZ8J | 99 | 409808 | 356320 |
| ATWGJ53DEL | NA | 611014 | 85770 |
| AT0TFNDWZG | 93 | 350110 | 203864 |
| AT6PMLSMAT | 71 | 677646 | 118692 |
| ATBSEDOKCN | 93 | 452264 | 293360 |
| ATAU1LDLE2 | NA | 624690 | 93593 |
| AT4FE9C8N9 | 84 | 577310 | 128734 |
| ATFY0YNZ33 | 84 | 686796 | 154397 |
| ATNGW65XYZ | 86 | 414068 | 315045 |
| AT3K7JLNGT | 14 | 758206 | 118737 |
| ATP3ZOJFI9 | 1 | 1039286 | 123049 |
| ATM6YA1R0Z | 99 | 507290 | 288831 |

| | | | |
|---|---|---|---|
| No_barcode_13 | 81 | 890958 | 246545 |
| ATNTXE6UZQ | 98 | 824780 | 384688 |
| ATUZ85D2VU | 99 | 682910 | 494599 |
| AT1W159O34 | 32 | 563916 | 109791 |
| ATP8WMDMZK | 100 | 858962 | 701834 |
| ATAJAL9JG4 | 98 | 654744 | 458966 |
| ATCZTKHDF4 | 100 | 783938 | 658101 |
| ATP2AGQNK1 | 77 | 517578 | 308320 |
| ATTYEJSDWY | 100 | 462752 | 374615 |
| ATSMWG98PK | 100 | 293600 | 252912 |
| ATCRC6FGN5 | NA | 2 | NA |
| ATG0HMTYYE | 99 | 528428 | 372524 |
| No barcode 2 | 93 | 851948 | 300344 |
| ATGLSXIQIT | NA | 2 | NA |
| ATBH4IK27M | 99 | 460436 | 284508 |
| ATA5WLTAK0 | 97 | 388442 | 338390 |
| ATV6AW7859 | 99 | 530482 | 424853 |
| ATACET5NJ8 | 99 | 686048 | 262110 |
| ATR9JWB9VK | NA | NA | NA |
| ATMYVE1QEK | 96 | 601906 | 213699 |
| ATNB4UEUAK | 98 | 691016 | 475180 |
| ATKR3E8K57 | 99 | 366958 | 304051 |
| ATEHBAOC25 | 100 | 358936 | 309877 |
| ATG75FDZ90 | 97 | 345068 | 270151 |
| ATXXSAVO9F | 99 | 454890 | 386104 |
| ATRX649RNA | 98 | 349086 | 148045 |
| ATS90Q5O1U | 85 | 612284 | 128288 |
| ATBW04JJJ9 | 99 | 415074 | 343636 |
| AT5M1NUPQS | 99 | 369402 | 309504 |
| ATWY6KKEA5 | 63 | 616968 | 112531 |
| ATQN1DOYOX | 95 | 366642 | 285513 |
| ATJUCHFV8Z | 99 | 518570 | 295505 |
| ATZYNED0C2 | 99 | 492508 | 394212 |
| ATCVEU14QN | 80 | 785162 | 155348 |
| ATZBGZQ7GK | 42 | 553134 | 98761 |
| ATALAP7D6L | 91 | 525094 | 211252 |
| No barcode 4 | 97 | 559098 | 354832 |
| ATFDG7UFZM | 85 | 468376 | 139461 |
| ATTVOS73K4 | 99 | 494720 | 395135 |

| | | | |
|---|---|---|---|
| AT9E5WUVE3 | 99 | 607072 | 307272 |
| AT0U00ZFF4 | 59 | 761988 | 160234 |
| ATPDK4X5OH | 94 | 398224 | 312593 |
| ATTYI7D5JH | 92 | 317232 | 224871 |
| AT8DP3WGHB | 95 | 373098 | 308633 |
| ATHLVVS4Y4 | NA | 753466 | 124774 |
| ATM53BCTFV | 93 | 523742 | 202461 |
| ATLOX1CQAQ | 79 | 544806 | 175590 |
| ATPVOTRGYM | 99 | 172966 | 146251 |
| AT0RH4DFUV | 93 | 573020 | 480617 |
| ATZ0NRXDCK | 95 | 206960 | 178578 |
| ATGUE1WYC7 | 53 | 493418 | 176869 |
| ATCZIZF68Z | 74 | 582974 | 176985 |
| ATNTDGQ0ZV | 96 | 306770 | 254238 |
| ATBZPSGVAB | 99 | 329430 | 291826 |
| ATJ0KWF0TS | 98 | 378290 | 168951 |
| ATWJ2D0980 | 98 | 382652 | 301589 |
| ATMDAY79PQ | 86 | 513622 | 260852 |
| ATCXJM1DBO | 96 | 419218 | 289501 |
| ATKJWPMFO2 | 76 | 403958 | 204195 |
| NO BARCODE "TIZZY" | 92 | 178138 | 162858 |
| ATHJWSM3HA | 78 | 508560 | 157250 |
| AT3ZC8GJPW | 99 | 344554 | 306681 |
| ATFGKWPAZL | 95 | 383338 | 319075 |
| ATFX9LZCHR | 99 | 357588 | 288248 |
| AT4VBM7WQH | 97 | 490514 | 377684 |
| AT96YQ9U13 | 99 | 382846 | 316223 |
| AT4J8ARPEG | 99 | 266736 | 228946 |
| No Barcode 6 | 81 | 562934 | 273771 |
| ATJR9OSFWD | 99 | 651960 | 287733 |
| AT7FWDGTYF | 55 | 717400 | 190486 |
| ATWYRUEAD7 | 47 | 628916 | 126046 |
| ATPLL5N312 | 67 | 577476 | 194399 |
| AT7MY3T1E9 | NA | NA | NA |
| No Barcode 5 BN | 95 | 536304 | 384937 |
| No Barcode 5 ihealth | 99 | 421328 | 365889 |
| ATUKMG6K7J | 98 | 463306 | 295424 |
| ATZBGZQ7GK-2 | 10 | 782986 | 84241 |

| | | | |
|---|---|---|---|
| ATPXOOQ032 | 91 | 400474 | 296893 |
| ATPL7GXV96 | 76 | 891358 | 275845 |
| ATPWR5IE4D | 100 | 433216 | 384950 |
| ATZ8EZY7IR | 94 | 550374 | 299563 |
| AT1CHWU504 | 5 | 995852 | 159976 |
| ATQOATMEB2 | 69 | 669062 | 87444 |
| No BARCODE 7 | 96 | 234600 | 207717 |
| ATEZEING5R | 98 | 525892 | 470022 |
| ATPAVSLTVZ | 98 | 515332 | 447870 |
| AT0ERFB1HF | 99 | 401720 | 346009 |
| ATSAO2XRUQ-2 | 98 | 608670 | 532227 |
| ATZJS41EVX | 98 | 574852 | 337597 |
| AT781XIB54 | 98 | 669908 | 541534 |
| ATPDSVRWH9 | 97 | 527216 | 455656 |
| AT0AWTMMKP | 96 | 492222 | 428600 |
| ATSAO2XRUQ | 96 | 479342 | 351492 |
| ATOI9WCZ81 | 99 | 743776 | 505064 |
| ATDDTZIOII | 97 | 680874 | 513429 |
| ATO4IQDBLG | 99 | 519930 | 423851 |
| AT76LFJKEJ | 95 | 724068 | 544041 |
| AT7LKS02ZY | 98 | 661996 | 504998 |
| ATMBVXFJQN | 98 | 514408 | 403974 |
| ATHVKKGPO2 | 99 | 576912 | 313256 |
| ATHX4ESJMU | 99 | 363530 | 209804 |
| AT4G966SRR | 98 | 727058 | 580634 |
| AT4J6GKZJV | 96 | 642310 | 347757 |
| AT4C6YKBZZ | 96 | 554572 | 387180 |
| ATOMINHMVO | 99 | 641806 | 262231 |
| AT7H83A7O2 | 90 | 586554 | 283807 |
| ATRVT2XAYT | 95 | 682452 | 469014 |
| ATODO9Q9H0 | 95 | 486940 | 292397 |
| AT05KGLKQ6 | 93 | 618334 | 226536 |
| ATN3EBDCSD | 92 | 610662 | 259921 |
| ATWXMLXGNP | 99 | 798352 | 300537 |
| ATJ92HKIPP | 44 | 961196 | 207289 |
| ATBSWKRXY3 | 86 | 948954 | 321611 |
| ATEEO64Q5W | 87 | 688572 | 253601 |
| AT25WIBJFT | 94 | 938212 | 255607 |
| AT5QBZRJ2P | 86 | 750942 | 337419 |

| | | | |
|---|---|---|---|
| ATKOAUDDAH-2 | 99 | 517210 | 337659 |
| ATU0HRPQER | 56 | 569040 | 99157 |
| ATUB1RTN5L | 97 | 430372 | 377749 |
| ATXI9FLPF9 | 34 | 873854 | 167279 |
| AT32O2dczr | NA | 10 | 2 |
| AT5CGJV0G0 | NA | 58 | 16 |
| Ath33n84qi | NA | 6 | 4 |
| ATKC3UWIMM | NA | 22 | 2 |
| ATKOAUDDAH | NA | 8 | NA |
| ATXJTJUN2K | NA | 54 | 44 |
| ATZPCYXSJB | NA | 22 | 14 |
| No_barcode_14 | 94 | 636822 | 369131 |
| No barcode 8 | 97 | 760660 | 370204 |
| No barcode 9 | 80 | 183516 | 51627 |
| 30147_AT0KKXXXTO_01 | 82 | 1352878 | 336231 |
| 30147_AT1F559LBQ_01 | 81 | 1046026 | 333562 |
| 30147_AT2T3FKNIS_01 | 98 | 812156 | 673725 |
| 30147_AT61VLINZO_01 | 71 | 922942 | 277725 |
| 30147_AT67EY0KZD_01 | 73 | 950004 | 230564 |
| 30147_AT7M9SP0UD_01 | 94 | 840602 | 633794 |
| 30147_AT85DBPO8X_01 | 89 | 976366 | 405103 |
| 30147_ATEJ3VUYVU_01 | 94 | 440728 | 393406 |
| 30147_ATF1KO758R_01 | 87 | 908078 | 251562 |
| 30147_ATH9QR4BYN_01 | 97 | 729502 | 645676 |
| 30147_ATIHCP9JIH_01 | 93 | 705846 | 613468 |
| 30147_ATKKB24DOT_01 | 88 | 516452 | 445004 |
| 30147_ATLMMUI9VT_01 | 68 | 942500 | 204324 |
| 30147_ATM2N85W29_01 | 90 | 1166916 | 683261 |
| 30147_ATMRYCFWRL_01 | 94 | 1069858 | 592132 |
| 30147_ATN7ZZTTSN_01 | 90 | 509106 | 281025 |
| 30147_ATPHZDSO0Y_01 | 87 | 868694 | 239250 |
| 30147_ATR1EVNFS3_01 | 82 | 1009044 | 739865 |
| 30147_ATRGK0DY0P_01 | 44 | 1218360 | 224435 |
| 30147_ATTG78KYMH_01 | 76 | 1161658 | 410103 |
| 30147_ATTKH7F78W_01 | 67 | 1018114 | 269172 |
| 30147_ATTX5ZVWSC_01 | 2 | 1396058 | 243182 |
| 30147_ATVRGVTXY7_01 | 4 | 1325366 | 253608 |
| 30147_ATWB8820OC_01 | 93 | 617134 | 535170 |
| 30147_ATWI85964T_01 | 78 | 1351490 | 296960 |

| | | | |
|---|---|---|---|
| 30147_ATXQQQFFSL_01 | 84 | 672456 | 571298 |
| 30147_ATXY5LQ12H_01 | 99 | 977534 | 461780 |
| 30147_ATY86E0TCY_01 | 88 | 1321354 | 319201 |
| 30147_ATYIMJ1U3N_01 | 98 | 577478 | 208854 |
| 30147_ATZ2JDFJD1_01 | 92 | 627986 | 294178 |

**Supplementary Table 2. List of accession numbers for Genbank and SRA and virus names for GI-SAID to access the rapid antigen test sequences generated as part of our library rapid antigen test community surveillance program**

| Internal ID | SRA accession number | Genbank accession number | GISAID virus name |
|---|---|---|---|
| ATX2RLNE3M_2 | SAMN40752681 | PP761650 | hCoV-19/USA/WI-UW-15394/2023 |
| ATAC3S9AX4_2 | SAMN40752680 | PP761651 | hCoV-19/USA/WI-UW-15393/2023 |
| AT25TA545F_2 | SAMN40752679 | PP761655 | hCoV-19/USA/WI-UW-15392/2023 |
| ATKCPI0YMU_2 | SAMN40752678 | PP761653 | hCoV-19/USA/WI-UW-15391/2023 |
| ATRUPA5DJ5_2 | SAMN40752677 | | hCoV-19/USA/WI-UW-15390/2023 |
| ATGRM0X3NG_2 | SAMN40752676 | PP761657 | hCoV-19/USA/WI-UW-15389/2023 |
| ATQC4UXFHL_2 | SAMN40752675 | PP761658 | hCoV-19/USA/WI-UW-15388/2023 |
| ATX2RLNE3M_1 | SAMN40752674 | PP761649 | hCoV-19/USA/WI-UW-15387/2023 |
| ATAC3S9AX4_1 | SAMN40752673 | PP761647 | hCoV-19/USA/WI-UW-15386/2023 |
| AT25TA545F_1 | SAMN40752672 | PP761652 | hCoV-19/USA/WI-UW-15385/2023 |
| ATKCPI0YMU_1 | SAMN40752671 | PP761659 | hCoV-19/USA/WI-UW-15384/2023 |
| ATRUPA5DJ5_1 | SAMN40752670 | | hCoV-19/USA/WI-UW-15383/2023 |
| ATFFPBDGC7_1 | SAMN40752669 | PP761648 | hCoV-19/USA/WI-UW-15382/2023 |
| ATGRM0X3NG_1 | SAMN40752668 | PP761654 | hCoV-19/USA/WI-UW-15381/2023 |
| ATQC4UXFHL_1 | SAMN40752667 | PP761656 | hCoV-19/USA/WI-UW-15380/2023 |
| ATM6YA1R0Z | SAMN40752666 | PP747776 | hCoV-19/USA/WI-UW-15517/2023 |
| No-barcode-8 | SAMN40752665 | PP747769 | hCoV-19/USA/WI-UW-15516/2024 |
| No-barcode-7 | SAMN40752664 | PP747798 | hCoV-19/USA/WI-UW-15515/2024 |
| ATWXMLXGNP | SAMN40752663 | PP747708 | hCoV-19/USA/WI-UW-15514/2024 |
| ATUB1RTN5L | SAMN40752662 | PP747774 | hCoV-19/USA/WI-UW-15513/2024 |
| ATSAO2XRUQ-2 | SAMN40752661 | PP747705 | hCoV-19/USA/WI-UW-15512/2024 |
| ATSAO2XRUQ-1 | SAMN40752660 | PP747696 | hCoV-19/USA/WI-UW-15511/2024 |
| ATRVT2XAYT | SAMN40752659 | PP747764 | hCoV-19/USA/WI-UW-15510/2024 |
| ATPDSVRWH9 | SAMN40752658 | PP747781 | hCoV-19/USA/WI-UW-15509/2024 |
| ATPAVSLTVZ | SAMN40752657 | PP747743 | hCoV-19/USA/WI-UW-15508/2024 |
| ATOMINHMVO | SAMN40752656 | PP747702 | hCoV-19/USA/WI-UW-15507/2024 |
| ATOI9WCZ81 | SAMN40752655 | PP747775 | hCoV-19/USA/WI-UW-15506/2024 |
| ATODO9Q9H0 | SAMN40752654 | PP747730 | hCoV-19/USA/WI-UW-15505/2024 |
| ATO4IQDBLG | SAMN40752653 | PP747734 | hCoV-19/USA/WI-UW-15504/2024 |
| ATN3EBDCSD | SAMN40752652 | PP747733 | hCoV-19/USA/WI-UW-15503/2024 |
| ATMBVXFJQN | SAMN40752651 | PP747750 | hCoV-19/USA/WI-UW-15502/2024 |
| ATKOAUDDAH-2 | SAMN40752650 | PP747765 | hCoV-19/USA/WI-UW-15501/2024 |
| ATHX4ESJMU | SAMN40752649 | PP747738 | hCoV-19/USA/WI-UW-15500/2024 |

| | | | |
|---|---|---|---|
| ATHVKKGPO2 | SAMN40752648 | PP747700 | hCoV-19/USA/WI-UW-15499/2024 |
| ATEZEING5R | SAMN40752647 | PP747744 | hCoV-19/USA/WI-UW-15498/2024 |
| ATDDTZIOII | SAMN40752646 | PP747740 | hCoV-19/USA/WI-UW-15497/2024 |
| AT7LKS02ZY | SAMN40752645 | PP747770 | hCoV-19/USA/WI-UW-15496/2024 |
| AT7H83A7O2 | SAMN40752644 | PP747711 | hCoV-19/USA/WI-UW-15495/2024 |
| AT781XIB54 | SAMN40752643 | PP747701 | hCoV-19/USA/WI-UW-15494/2024 |
| AT76LFJKEJ | SAMN40752642 | PP747723 | hCoV-19/USA/WI-UW-15493/2024 |
| AT4J6GKZJV | SAMN40752641 | PP747693 | hCoV-19/USA/WI-UW-15492/2024 |
| AT4G966SRR | SAMN40752640 | PP747749 | hCoV-19/USA/WI-UW-15491/2024 |
| AT4C6YKBZZ | SAMN40752639 | PP747793 | hCoV-19/USA/WI-UW-15490/2024 |
| AT25WIBJFT | SAMN40752638 | PP747697 | hCoV-19/USA/WI-UW-15489/2024 |
| AT0ERFB1HF | SAMN40752637 | PP747698 | hCoV-19/USA/WI-UW-15488/2024 |
| AT0AWTMMKP | SAMN40752636 | PP747760 | hCoV-19/USA/WI-UW-15487/2024 |
| AT05KGLKQ6 | SAMN40752635 | PP747692 | hCoV-19/USA/WI-UW-15486/2024 |
| ATFGKWPAZL | SAMN40752634 | PP747796 | hCoV-19/USA/WI-UW-15485/2023 |
| NO-BARCODE-TIZZY | SAMN40752633 | PP747762 | hCoV-19/USA/WI-UW-15484/2023 |
| No-BARCODE-7 | SAMN40752632 | PP747803 | hCoV-19/USA/WI-UW-15483/2023 |
| No-Barcode-5-ihealth | SAMN40752631 | PP747707 | hCoV-19/USA/WI-UW-15482/2023 |
| No-Barcode-5-BN | SAMN40752630 | PP747800 | hCoV-19/USA/WI-UW-15481/2023 |
| ATPXOOQ032 | SAMN40752629 | PP747728 | hCoV-19/USA/WI-UW-15480/2023 |
| ATZ8EZY7IR | SAMN40752628 | PP747782 | hCoV-19/USA/WI-UW-15479/2023 |
| ATZ0NRXDCK | SAMN40752627 | PP747789 | hCoV-19/USA/WI-UW-15478/2023 |
| ATWJ2D0980 | SAMN40752626 | PP747742 | hCoV-19/USA/WI-UW-15477/2023 |
| ATUKMG6K7J | SAMN40752625 | PP747785 | hCoV-19/USA/WI-UW-15476/2023 |
| ATPWR5IE4D | SAMN40752624 | PP747695 | hCoV-19/USA/WI-UW-15475/2023 |
| ATPVOTRGYM | SAMN40752623 | PP747802 | hCoV-19/USA/WI-UW-15474/2023 |
| ATNTDGQ0ZV | SAMN40752622 | PP747778 | hCoV-19/USA/WI-UW-15473/2023 |
| ATM53BCTFV | SAMN40752621 | PP747737 | hCoV-19/USA/WI-UW-15472/2023 |
| ATJR9OSFWD | SAMN40752620 | PP747792 | hCoV-19/USA/WI-UW-15471/2023 |
| ATJ0KWF0TS | SAMN40752619 | PP747739 | hCoV-19/USA/WI-UW-15470/2023 |
| ATFX9LZCHR | SAMN40752618 | PP747713 | hCoV-19/USA/WI-UW-15469/2023 |
| ATCXJM1DBO | SAMN40752617 | PP747790 | hCoV-19/USA/WI-UW-15468/2023 |
| ATBZPSGVAB | SAMN40752616 | PP747795 | hCoV-19/USA/WI-UW-15467/2023 |
| AT96YQ9U13 | SAMN40752615 | PP747791 | hCoV-19/USA/WI-UW-15466/2023 |
| AT4VBM7WQH | SAMN40752614 | PP747772 | hCoV-19/USA/WI-UW-15465/2023 |
| AT4J8ARPEG | SAMN40752613 | PP747721 | hCoV-19/USA/WI-UW-15464/2023 |
| AT3ZC8GJPW | SAMN40752612 | PP747767 | hCoV-19/USA/WI-UW-15463/2023 |
| AT0RH4DFUV | SAMN40752611 | PP747779 | hCoV-19/USA/WI-UW-15462/2023 |
| No-barcode-4 | SAMN40752610 | PP747735 | hCoV-19/USA/WI-UW-15461/2023 |
| No-barcode-2 | SAMN40752609 | PP747717 | hCoV-19/USA/WI-UW-15460/2023 |
| ATZYNED0C2 | SAMN40752608 | PP747766 | hCoV-19/USA/WI-UW-15459/2023 |
| ATXXSAVO9F | SAMN40752607 | PP747741 | hCoV-19/USA/WI-UW-15458/2023 |
| ATV6AW7859 | SAMN40752606 | PP747727 | hCoV-19/USA/WI-UW-15457/2023 |
| ATUZ85D2VU | SAMN40752605 | PP747722 | hCoV-19/USA/WI-UW-15456/2023 |
| ATTYI7D5JH | SAMN40752604 | PP747745 | hCoV-19/USA/WI-UW-15455/2023 |

| | | | |
|---|---|---|---|
| ATTYEJSDWY | SAMN40752603 | PP747703 | hCoV-19/USA/WI-UW-15454/2023 |
| ATTVOS73K4 | SAMN40752602 | PP747694 | hCoV-19/USA/WI-UW-15453/2023 |
| ATSMWG98PK | SAMN40752601 | PP747773 | hCoV-19/USA/WI-UW-15452/2023 |
| ATRX649RNA | SAMN40752600 | PP747783 | hCoV-19/USA/WI-UW-15451/2023 |
| ATQN1DOYOX | SAMN40752599 | PP747746 | hCoV-19/USA/WI-UW-15450/2023 |
| ATPDK4X5OH | SAMN40752598 | PP747801 | hCoV-19/USA/WI-UW-15449/2023 |
| ATP8WMDMZK | SAMN40752597 | PP747788 | hCoV-19/USA/WI-UW-15448/2023 |
| ATNTXE6UZQ | SAMN40752596 | PP747756 | hCoV-19/USA/WI-UW-15447/2023 |
| ATNB4UEUAK | SAMN40752595 | PP747709 | hCoV-19/USA/WI-UW-15446/2023 |
| ATMYVE1QEK | SAMN40752594 | PP747758 | hCoV-19/USA/WI-UW-15445/2023 |
| ATKR3E8K57 | SAMN40752593 | PP747768 | hCoV-19/USA/WI-UW-15444/2023 |
| ATJUCHFV8Z | SAMN40752592 | PP747799 | hCoV-19/USA/WI-UW-15443/2023 |
| ATG75FDZ90 | SAMN40752591 | PP747780 | hCoV-19/USA/WI-UW-15442/2023 |
| ATG0HMTYYE | SAMN40752590 | PP747754 | hCoV-19/USA/WI-UW-15441/2023 |
| ATEHBAOC25 | SAMN40752589 | PP747747 | hCoV-19/USA/WI-UW-15440/2023 |
| ATCZTKHDF4 | SAMN40752588 | PP747718 | hCoV-19/USA/WI-UW-15439/2023 |
| ATBW04JJJ9 | SAMN40752587 | PP747753 | hCoV-19/USA/WI-UW-15438/2023 |
| ATBH4IK27M | SAMN40752586 | PP747716 | hCoV-19/USA/WI-UW-15437/2023 |
| ATALAP7D6L | SAMN40752585 | PP747704 | hCoV-19/USA/WI-UW-15436/2023 |
| ATAJAL9JG4 | SAMN40752584 | PP747712 | hCoV-19/USA/WI-UW-15435/2023 |
| ATACET5NJ8 | SAMN40752583 | PP747763 | hCoV-19/USA/WI-UW-15434/2023 |
| ATA5WLTAK0 | SAMN40752582 | PP747757 | hCoV-19/USA/WI-UW-15433/2023 |
| AT9E5WUVE3 | SAMN40752581 | PP747751 | hCoV-19/USA/WI-UW-15432/2023 |
| AT8DP3WGHB | SAMN40752580 | PP747706 | hCoV-19/USA/WI-UW-15431/2023 |
| AT5M1NUPQS | SAMN40752579 | PP747719 | hCoV-19/USA/WI-UW-15430/2023 |
| AT1REAXVVZ | SAMN40752578 | PP747804 | hCoV-19/USA/WI-UW-15429/2023 |
| AT9M50GXL5 | SAMN40752577 | PP747784 | hCoV-19/USA/WI-UW-15428/2023 |
| AT7V74R0RW | SAMN40752576 | PP747715 | hCoV-19/USA/WI-UW-15427/2023 |
| ATHXJM6OTS | SAMN40752575 | PP747794 | hCoV-19/USA/WI-UW-15426/2023 |
| AT7ZH839N8 | SAMN40752574 | PP747786 | hCoV-19/USA/WI-UW-15425/2023 |
| AT8X5RUNJL | SAMN40752573 | PP747710 | hCoV-19/USA/WI-UW-15424/2023 |
| ATRBJD97CM | SAMN40752572 | PP747736 | hCoV-19/USA/WI-UW-15423/2023 |
| ATF5O133LQ | SAMN40752571 | PP747726 | hCoV-19/USA/WI-UW-15422/2023 |
| ATUQ7230RN | SAMN40752570 | PP747731 | hCoV-19/USA/WI-UW-15421/2023 |
| AT0TFNDWZG | SAMN40752569 | PP747724 | hCoV-19/USA/WI-UW-15420/2023 |
| ATCLJ5EIQG | SAMN40752568 | PP747725 | hCoV-19/USA/WI-UW-15419/2023 |
| AT5UTIINHL | SAMN40752567 | | hCoV-19/USA/WI-UW-15418/2023 |
| AT06HSAZ8J | SAMN40752566 | PP747771 | hCoV-19/USA/WI-UW-15417/2023 |
| ATBSEDOKCN | SAMN40752565 | PP747761 | hCoV-19/USA/WI-UW-15416/2023 |
| ATGRM0X3NG | SAMN40752564 | PP747797 | hCoV-19/USA/WI-UW-15415/2023 |
| ATQC4UXFHL | SAMN40752563 | PP747720 | hCoV-19/USA/WI-UW-15414/2023 |
| ATU2UEZGUU | SAMN40752562 | PP747755 | hCoV-19/USA/WI-UW-15413/2023 |
| ATKCPI0YMU | SAMN40752561 | PP747787 | hCoV-19/USA/WI-UW-15412/2023 |
| ATRUPA5DJ5 | SAMN40752560 | PP747732 | hCoV-19/USA/WI-UW-15411/2023 |
| ATFFPBDGC7 | SAMN40752559 | PP747777 | hCoV-19/USA/WI-UW-15410/2023 |

| | | | |
|---|---|---|---|
| AT25TA545F | SAMN40752558 | PP747729 | hCoV-19/USA/WI-UW-15409/2023 |
| ATX2RLNE3M | SAMN40752557 | PP747699 | hCoV-19/USA/WI-UW-15408/2023 |
| ATAC3S9AX4 | SAMN40752556 | PP747759 | hCoV-19/USA/WI-UW-15407/2023 |
| Lib-RAT-5-CC | SAMN40752555 | PP747752 | hCoV-19/USA/WI-UW-15406/2023 |
| Lib-RAT-4-CC | SAMN40752554 | PP747748 | hCoV-19/USA/WI-UW-15405/2023 |
| Lib-RAT-2-CC | SAMN40752553 | PP747691 | hCoV-19/USA/WI-UW-15404/2023 |
| Lib-RAT-10-CC | SAMN40752552 | PP747714 | hCoV-19/USA/WI-UW-15403/2023 |
